# Antibiotic pharmacokinetics in infected pleural effusions

**DOI:** 10.1101/2023.09.08.23295282

**Authors:** David T Arnold, Liam Read, Oliver Waddington, Fergus W Hamilton, Sonia Patole, Jessica Hughes, Alice Milne, Alan Noel, Mark Bayliss, Nick A Maskell, Alasdair MacGowan

## Abstract

Pleural infection is usually treated with empirical broad-spectrum antibiotics but limited data exists around their penetrance into the infected pleural space. We performed a pharmacokinetic study analysing the concentration of 5 intravenous antibiotics across 146 separate timepoints in 35 patients (amoxicillin, metronidazole, piperacillin-tazobactam, clindamycin and cotrimoxazole). All antibiotics tested, apart from co-trimoxazole, reach pleural fluid levels equivalent to levels within blood, and well above the relevant minimum inhibitory concentrations. The results demonstrate that concerns around the penetration of commonly used antibiotics, apart from co-trimoxazole, into the infected pleural space are unfounded.

## Introduction

Pleural infection is a serious clinical condition with a mortality of up to 15%[1]. Patients spend extended periods in hospital for pleural fluid drainage and intravenous antibiotic administration[1, 2]. There has been concern around the ability of antibiotics to reach therapeutic levels at the site of infection[2].

## Methods

We recruited patients presenting to a single UK tertiary pleural centre with pleural infection. Patients were consented for regular pleural fluid sampling (via chest tube) with synchronous blood sampling timed with antibiotic administration (1, 2, 4, 6 hours post dose for a six-hourly (QDS) regimen; 1, 4, 8 hours for an 8-hourly (TDS) regimen; 1, 4, 8 and 12 hourly (BD) for a 12-hourly regimen). The study received ethical approval from the East of Scotland Research Ethics Service (REC reference:19/ES/0075).

Analytes were assayed using liquid chromatography mass spectrometry assays (LC-MS/MS). Deproteinised samples were injected into a Shimadzu LC system and gradient separation performed on a 50 × 2.1 mm ID 2.6 μm Kinetex XBC18 column using mobile phases containing water/acetonitrile/formic acid (0.1%,v/v). Analytes were selectively detected using a positive ion electrospray source on AB Sciex 4000 QTrap MS. Concentrations in test samples were calculated by Sciex Analyst™ software using calibrators prepared in the same biological matrix.

Total drug concentrations were plotted with area under the curve (AUC) from time zero to next dose determined by the trapezoidal rule. The penetration ratio (PR) for each patient was obtained by dividing the AUC for pleural fluid by the AUC for blood. Pearson correlation coefficient was used to assess the correlation between antibiotic concentrations and pleural fluid pH. A paired T-test was used to compare concentrations between loculated and non-loculated effusions.

## Results

Thirty-five patients were recruited between October 2019 and March 2022. Six patients met the criteria for frank empyema with the others meeting at least one criterion for pleural infection [3]. There was a male predominance (77%) with a median age of 76 (IQR 65-79)). Full pharmacokinetic analysis could be performed for the antibiotics shown in Table 1 with summary curves shown in Figure 1.

**Table 1:** Antibiotics assayed, dosing schedule and number of timepoints assessed.

| Antibiotic | Route | Dose (frequency) | Timepoints | Patients |
| --- | --- | --- | --- | --- |
| Amoxicillin | IV | 1000mg (8hr) | 44 | 10 |
| Metronidazole | IV | 500mg (8hr) | 31 | 8 |
| Piperacillin-tazobactam | IV | 4500mg (8hr) | 27 | 7 |
| Co-trimoxazole | IV | 960mg (12hr) | 24 | 5 |
| Clindamycin | IV | 600mg (6hr) | 20 | 5 |
| IV-Intravenous, mg-milligrams. |  |  |  |  |

**Figure 1:**
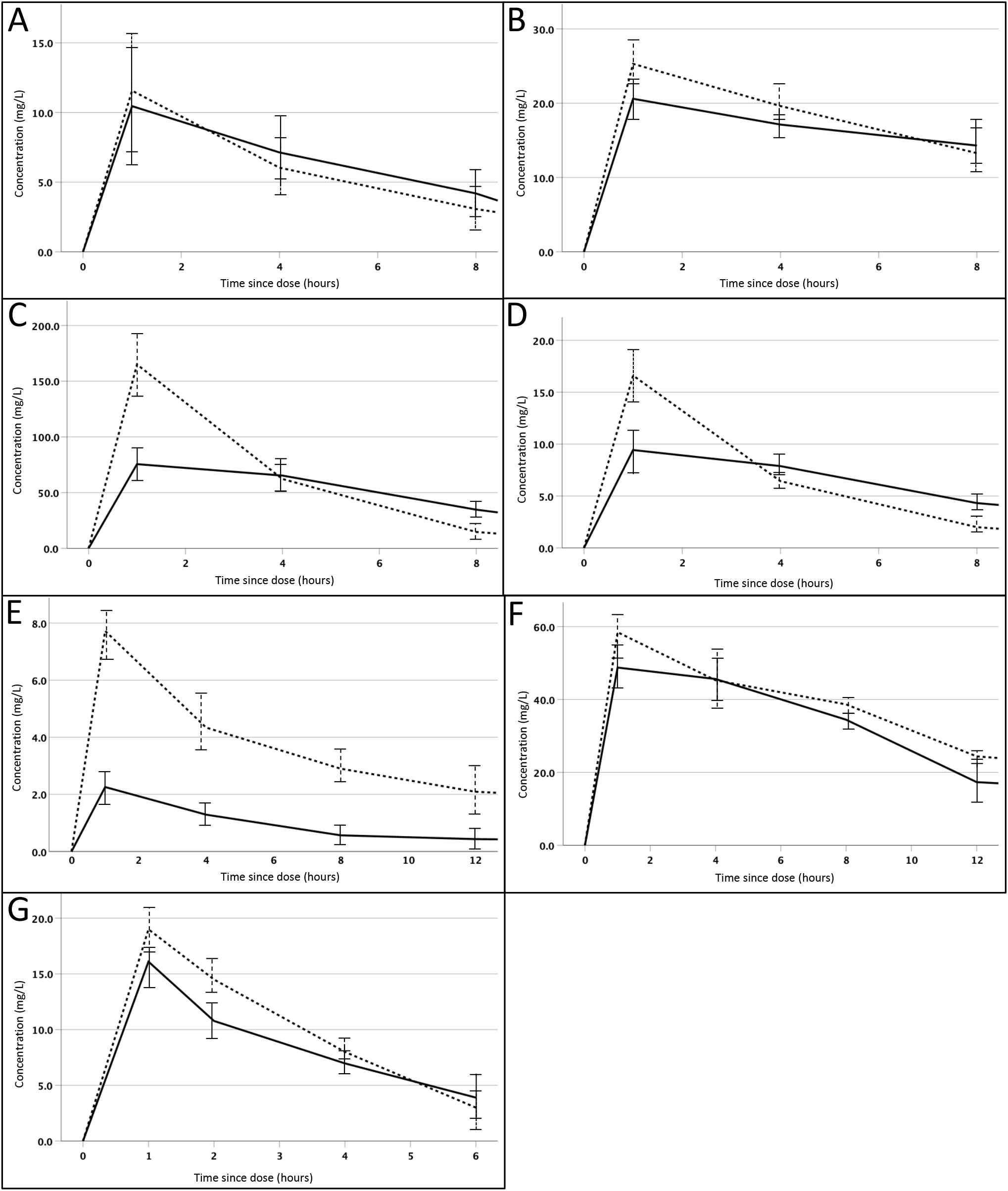
Mean (+/- standard deviation) intravenous antibiotic concentration-time curves in blood (dotted line) and pleural fluid (solid line): A- Amoxicillin, B- Metronidazole, C- Piperacillin (Piperacillin-Tazobactam), D- Tazobactam (Piperacillin-Tazobactam), E- Trimethoprim (Co-trimoxazole), F- Sulphamethoxazole (Co-trimoxazole), G- Clindamycin. Figure 1 footnote: MICs (EUCAST) of amoxicillin: Staph aureus 1mg/L, Haemophilus influenza 2mg/L, Streptococcus Milleri group 0.125mg/L, Streptococcus pneumoniae 0.06mg/L. MICs (EUCAST) of metronidazole: Prevotella 1mg/L, Gram positive anaerobes (class) 0.25mg/L, Fusobacterium 0.125mg/L. MICs (EUCAST) of piperacillin-tazobactam: Staph aureus 2mg/L, Haemophilus influenza 0.5mg/L, Streptococcus Milleri group 1mg/L, Escherichia coli 8mg/L, Streptococcus pneumoniae 0.064mg/L, MICs (EUCAST) of trimethoprim-sulfamethoxazole: Staph aureus 0.25mg/L, Haemophilus influenza 0.5mg/L, Streptococcus pneumoniae 1mg/L, Escherichia coli 0.5mg/L. MICs (EUCAST) of clindamycin: Staph aureus 0.25mg/L, Streptococcus Milleri group 0.25mg/L, Streptococcus pneumoniae 0.25mg/L. The MICs listed for common causative organisms are derived from the EUCAST website (https://www.eucast.org/mic_distributions_and_ecoffs/) which describes the MIC of ‘wild-type’ and therefore fully sensitive bacteria.

### Amoxicillin

The AUC_8hr_ for the dosing interval was marginally higher in pleural fluid compared to blood (62.8 vs 48.5, PR 1.1) and above the MIC of causative organisms for the entirety of the dosing schedule.

### Metronidazole

The AUC_8hr_ for metronidazole was similar between pleural fluid and blood (128.5 vs 151.5, PR 0.84).

### Piperacillin-Tazobactam

Piperacillin levels peaked rapidly in plasma but fell sharply at the 4-hour timepoint [4]. The AUC_8hr_ was similar between pleural fluid and blood (465 vs 648, PR 0.72). The AUC_8hr_ for tazobactam was similar comparing pleural fluid and blood (55.7 vs 59.5 respectively) with a ratio to piperacillin of 11.6%.

### Co-trimoxazole

The trimethoprim and sulfamethoxazole constituents of co-trimoxazole were measured separately. The AUC_12hr_ was much lower for trimethoprim in pleural fluid compared to blood (13.5 vs 47.2), with a PR of 0.29. The ratio for sulfamethoxazole was more equivalent (AUC_12hr_ 465.0 in blood vs 425.8 in pleural fluid, PR 0.92).

### Clindamycin

The AUC_6hr_ for clindamycin was similar between pleural fluid and blood (41.3 vs 51.3, PR 0.81).

### pH and loculation

Across the 7 compounds tested, there was no correlation between the mean concentration of antibiotic and the pH of the pleural fluid on sampling (p=0.74) or the presence of loculations within the effusion (p=0.51).

## Discussion

This is the largest study of antibiotic pharmacokinetics performed in infected pleural effusions. We have demonstrated that commonly used antibiotics of amoxicillin, metronidazole, piperacillin-tazobactam and clindamycin reach levels within the pleural fluid equivalent to that in blood and above the MIC for bacteria known to cause pleural infection. The trimethoprim element of co-trimoxazole did not reach the pleural fluid adequately, raising concerns about its use in established or developing pleural infection.

Despite antibiotic recommendations in guidelines and across study protocols, studies on the pleural penetration of many of these antibiotics have not been performed[5]. The most widely cited evidence on antibiotic concentrations in infected pleural spaces is from rabbit models of turpentine-induced pleural infection[6]. In 1987, Shohet demonstrated that gentamicin had diminished efficacy in pleural infection, leading to reduced aminoglycoside use for pleural infection[7]. Given stark differences in conclusions compared to human pharmacokinetic studies, results from animal models should be interpreted with caution[2].

Studies of antibiotic pharmacokinetics in human pleural infection are limited in sample size, methodology and antibiotic regimens. A review by Lau and colleagues reported that across the relevant 5 studies most antibiotics had been assessed following a single dose in between 1 to 3 patients with pleural infection. Thys et al measured aminoglycoside levels after a single intravenous dose in patients with uninfected versus infected purulent pleural infection (n=19 and n=11 respectively). In the infected effusions, levels were either undetectable or significantly lower than plasma, demonstrating the importance of performing pharmacokinetic studies using infected pleural fluid.

This study has demonstrated that for amoxicillin, metronidazole, piperacillin-tazobactam and clindamycin the pleural concentrations are equivalent to blood levels [6]. Pleural fluid trimethoprim levels were much lower than in blood across the dosing schedule. There is no previous literature on the pharmacokinetics of trimethoprim within infected pleural fluid. Extrapolating from other encapsulated infections there is evidence of poor penetration of co-trimoxazole into the gall bladder during acute cholecystitis [8]. When compared to the MIC for gram positives that commonly cause pleural infection, (e.g. *streptococcus pneumoniae* and the streptococcus virdans group) levels for trimethoprim were inadequate [9].

We were not able to perform sub-group analysis for individual antibiotics for factors such as patient age, fluid pH or degree of pleural loculation. Secondly, like many pharmacokinetic studies, the comparative MICs have been extrapolated from studies of wild-type bacteria as opposed to those with resistance patterns.

## Conclusion

The commonly used antibiotics amoxicillin, metronidazole, piperacillin-tazobactam and clindamycin reached levels equivalent to blood within infected pleural fluid. Low penetration of trimethoprim into the pleural space raised concerns about the use of cotrimoxazole for patients with pleural infection or parapneumonic effusions.

## Data Availability

Data available on request

## Acknowledgements

We would like to thank the patients who participated in this work.

